# Mean time to infection by small diffusing droplets containing SARS-CoV-2 during close social contacts

**DOI:** 10.1101/2021.04.01.21254802

**Authors:** U. Dobramysl, C. Sieben, D. Holcman

## Abstract

Airborne viruses such as SARS-CoV-2 are partly spreading through aerosols containing viral particles. Inhalation of infectious airborne particles can lead to infection, a route that can even be more predominant compared with droplet or contact transmission. To study the transmission between a susceptible and an infected person, we estimate the distribution of arrival times of small diffusing aerosol particles to the inhaled region located below the nose until the number of particles reaches a critical threshold. Our results suggest that although contamination by continuous respiration can take around 90 minutes at a distance of one meter, it is reduced to a few minutes when coughing or sneezing. Interestingly, there is not much differences between outdoors and indoors when the air is still. When a window is open inside an office, the infection time is reduced. Finally, wearing a mask leads to a delay in the time to infection. To conclude, diffusion analysis provides several key time scale of viral airborne transmission.

## 1 Introduction

Airborne SARS-CoV-2 viruses spread between humans in particular through respiratory droplets [1,2] expelled during breathing, coughing or sneezing from an infectious person. Droplets resulting from coughing or sneezing can infect a target person by entering into the respiratory mucosa. These droplets can be separated in coarse (> 5*μm*) and fine (≤ 5 *μm*) fractions. Although large particles can be respired and reach alveolar tissues [3], they usually fall to the ground quickly within one to two meters [4,5], However, smaller droplets (aerosols) remain suspended in the air for much longer periods of time, found to be infectious even after 12 hours [6], This route most likely leads to major spreading at home, work, hospitals and in urgent care medical units [7], Indeed, indoor space is a major SARS-CoV-2 infection risk [7] as the virus can be transmitted via speech droplets [8], Aerosol particles are able to travel hundreds of meters or more [9], Furthermore, fine particles were found to contain 8.8 times more viral copies than coarse particles [10], Yet, precisely quantifying the infectivity due to aerosols transmission remains difficult for several reasons: it is hard to measure the droplets despite recent efforts [8]; and the step from droplet dispersion to human contamination requires an assessment of the efficiency of infection. To get around this difficulties, data was obtained in Ferrets [11] although it still remained difficult to extrapolate these aerosol transmission measurements to human scale. In ferrets, sneezing increases the number of small droplets [11] and the majority of particles generated during sneezing were < 5*μm*. For example, exhaled aerosol particles from infected or naive ferrets during normal breathing (for 30 min) or sneezing (for 5 min) both lead to around 5000 droplets produced. In influenza, human study participants exhaled up to 20 virus RNA copies per minute [12], which is assumed to be of similar magnitude for SARS-CoV-2. The motion of aerosilized droplets is well approximated as a diffusion process, with a size-dependent diffusion coefficient ranging from *D* = 10^−4^*m*^2^/*s*, for a size around 0.1 *μm* to *D* = 10 ^−7^*m*^2^/*s* lor 5*μm* [13],

Here, we compute the time it takes for a susceptible person to get infected when in close proximity to an infected person. We explore how this transmission depends on the number of viral particles, air transportation parameters and the distance between two persons. We estimate the probability and the mean time of infection when the infectious and susceptible person are placed in the same room or outside using diffusion modeling and numerical simulations of aerosols. We evaluate the role of wearing a mask [14] in several different scenarios, including regular respiration cycles (exhalation) and coughing. While surgical masks reduce viral copy numbers in the coarse droplet fraction by a factor of 25, we focus here on the fine fraction, where they reduce the viral aerosol load only by a factor of 2.8 [10], Finally, we explore the sensitivity of several key parameters such as the diffusion coefficient or the viral copy number threshold required for infection. In summary, we found that talking to a infectious person for less than 100min does not lead to infection, while being exposed to a coughing person without protection can lead to infection within minutes. We conclude that wearing a mask can strongly suppress the propagation of infection via the small aerosol particle route.

## 2 Results

### 2.1 Viral propagation by small diffusing aerosol droplets: a diffusion modeling approach

We model viral diffusion and infection as follows: an infected person is positioned at a distance *d* from a susceptible person, either in a 3 x 2 x 2*m* room or outside (Fig.1A). We address the difference in the release of free diffusing aerosols during regular respiration (exhalation) and sneezing or coughing, by changing the initial distribution and increasing the number of droplets (see section 4.1): coughing is modeled by releasing 10^4^ viral particles instantaneously with a rate in a range between 30 seconds to 5 minutes [15], Each viral particle is contained in their own aerosolized droplet of 0.1*μ*m size [15,16], The full particle load is randomly and uniformly distributed in a sphere of 1*m* diameter directly in front of the infected person’s head.

**Figure 1:**
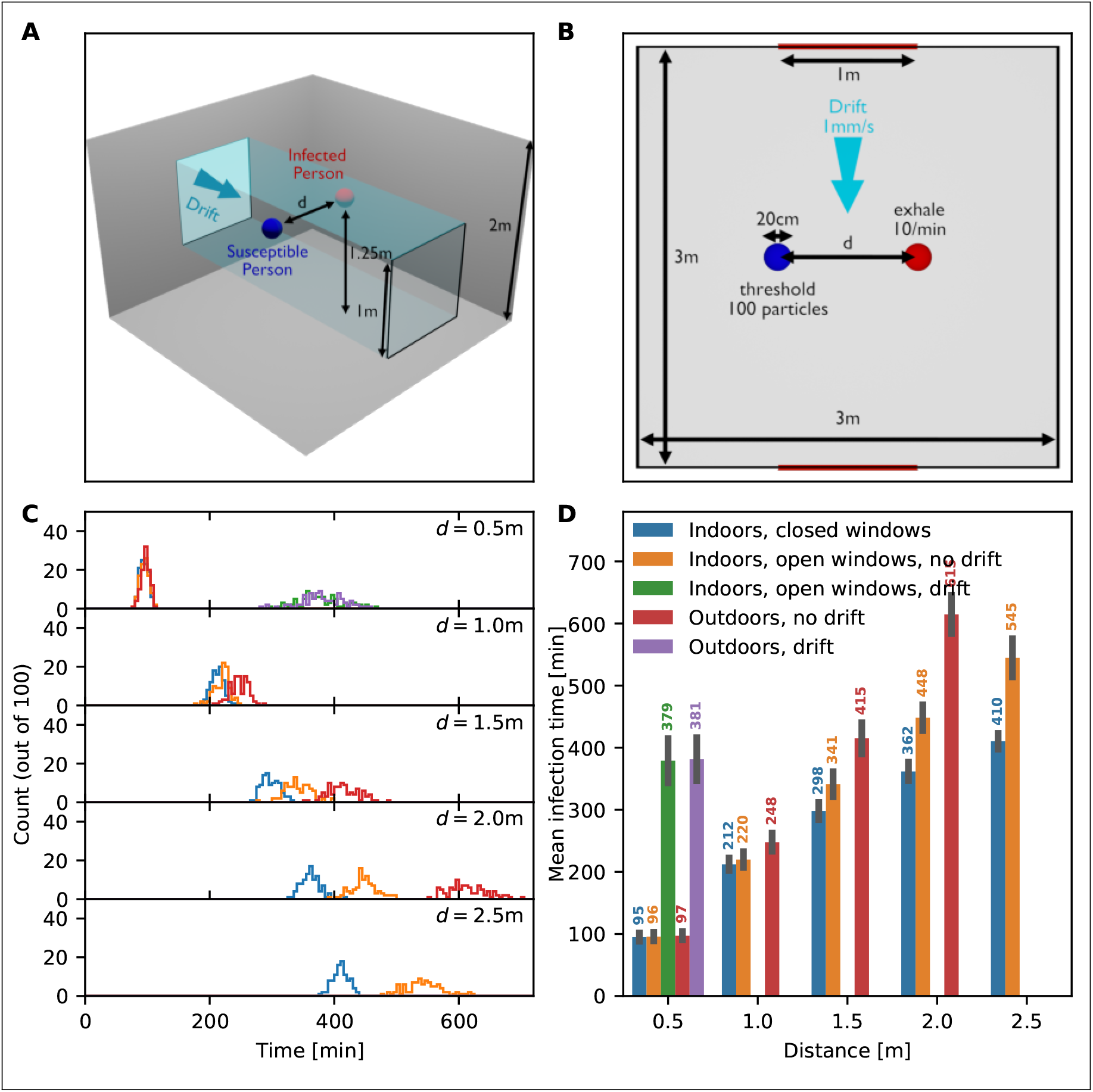
Simulations of infection by diffusing aerosols: Infection time distribution when an infected person is breathing only. (A) 3D schematic representation of a closed room containing an infected (red) and a susceptible (blue) person separated by a variable distance *d*. The room contains two windows which can be either open or closed, and a possible drift of air between the windows. (B) 2D schematic projection of (A). Room and window dimensions are displayed in the schematics. (C) Distribution of time of infection for distances *d* = 0.5,1,1.5, 2 and 2.5m and the following scenarios: Indoors with windows closed (blue), indoors with open windows (orange), indoors with open windows and a drift (green), outdoors without drift (red), and outdoors with drift (purple). (D) Mean infection time for the different scenarios. Missing bars indicate scenarios in which at least one realization had an infection time beyond 12 hours.

We also investigated breathing without coughing (in e.g. asymptomatic cases). In this case, we use a production of 10 particles per minute [16] at the location of the infected person’s head. The susceptible person is modeled as an absorbing sphere of 10cm radius around this person’s head. This accounts for the breathed-in volume of air during inhalation.

Infection occurs when a threshold number of 100 viral particles are inhaled by the susceptible person, accounting for the independent action hypothesis likely does not hold for SARS-CoV-2 [17],

In the indoors case, the room has a pair of windows in the centers of the long walls (opposite each other) that can either be open or closed. When the windows are open (where they act as absorbing surfaces), there can be a small draft (resulting in a drift of 1mm/s) in the volume between the windows or no draft. In the outdoors case, it is either wind-still, or the same drift is applied throughout the domain. Finally, the susceptible person can wear a mask, in which case only one in 2.8 particles is absorbed on average [10]).

### 2.2 Aerosol infection due to breathing indoors and outdoors

We first study the time to infection based on diffusing aerosols in a closed room, containing a susceptible person separated by a distance *d* from an infected person (Fig.1A-B). We run simulations to obtain the distribution of infection times to reach a cumulative infection threshold *T =* 100 of aerosol particles inhaled by the susceptible person (using 100 realizations). As we vary the distance *d* (Fig.1C), we find that the mean time of infection increases drastically: For *d* = 0.5,1 and 2*m*, we found a mean time of 95 ± 7 min, 212 ± 10 min and 362 ± 15 min (mean and standard deviation, respectively). Open windows alone, or even being outside, have no (for *d* = 0.5*m*) or only a small effect (*d* ≥ 1*m*) on these time scales (Fig. 1C-D). A small drift of 1mm/*s* perpendicular to the axis connecting the two persons leads to an increase of the time of infection to around 381 ± 35 minutes, a four-fold increase at a distance of *d* = 0.5*m*. For larger distances, we do not show the infection time because it is beyond our simulation time limit of 12 hours. Hence, in our model of diffusing aerosols generated by periodic breathing with 10 viral particles released per minute, we found that the infection threshold of 100 absorbed particles is reached in around 1.5 hours. Open windows without a drift does not change this time scale substantially, but ventilation in form of a drift does.

We derived the mathematical expression for the infection time as a function of distance (12) (see section 4.3). This provides a good fit to the simulation results for the outdoor case (Fig. S4).

We also looked at the importance of the direction of the air movement. Figure 1 shows the mean time to infection when air moves perpendicular to the line connecting the infected and the susceptible person. When the drift is applied parallel to this line (i.e. particles are blown from the infected person towards the susceptible person), the mean time to infection is reduced such that infection occurs at the half-hour mark even at a distance of *d* = 2.5*m* (Fig. S1). When the wind direction is antiparallel, infection is not occuring within any reasonable time frame (Fig. S1).

### 2.3 Coughing

We next evaluate the time to infection of a susceptible person using the same spatial configuration as before (Fig. 2A-B)). Here, the infected person coughs every 5 minutes. The time of infectious reduces drastically compared to breathing alone, as coughing leads to the instantaneous release of 10^4^ particles in front of the infected person. Our results show that the distribution for the times of infection are peaked around the mean: at a distance of *d =* 0.5*m*, the mean time to infection is 15 ± 1 minutes indoors and outdoors (regardless of the state of the windows) without a drift. This rises to 26 ± 2 minutes when there is a drift. At larger distances, a small drift more than triples the time to infection, which highlights the importance of distancing even when there is appropriate ventilation. Coughing at 30 second intervals shortens the time to infection by roughly a third (Fig. S2). Note that we assumed that the infected person is facing away from the susceptible person when coughing (light red shaded region in Fig. 1A-B). As before, we do not show the mean time to infection if it is larger than 12 hours.

**Figure 2:**
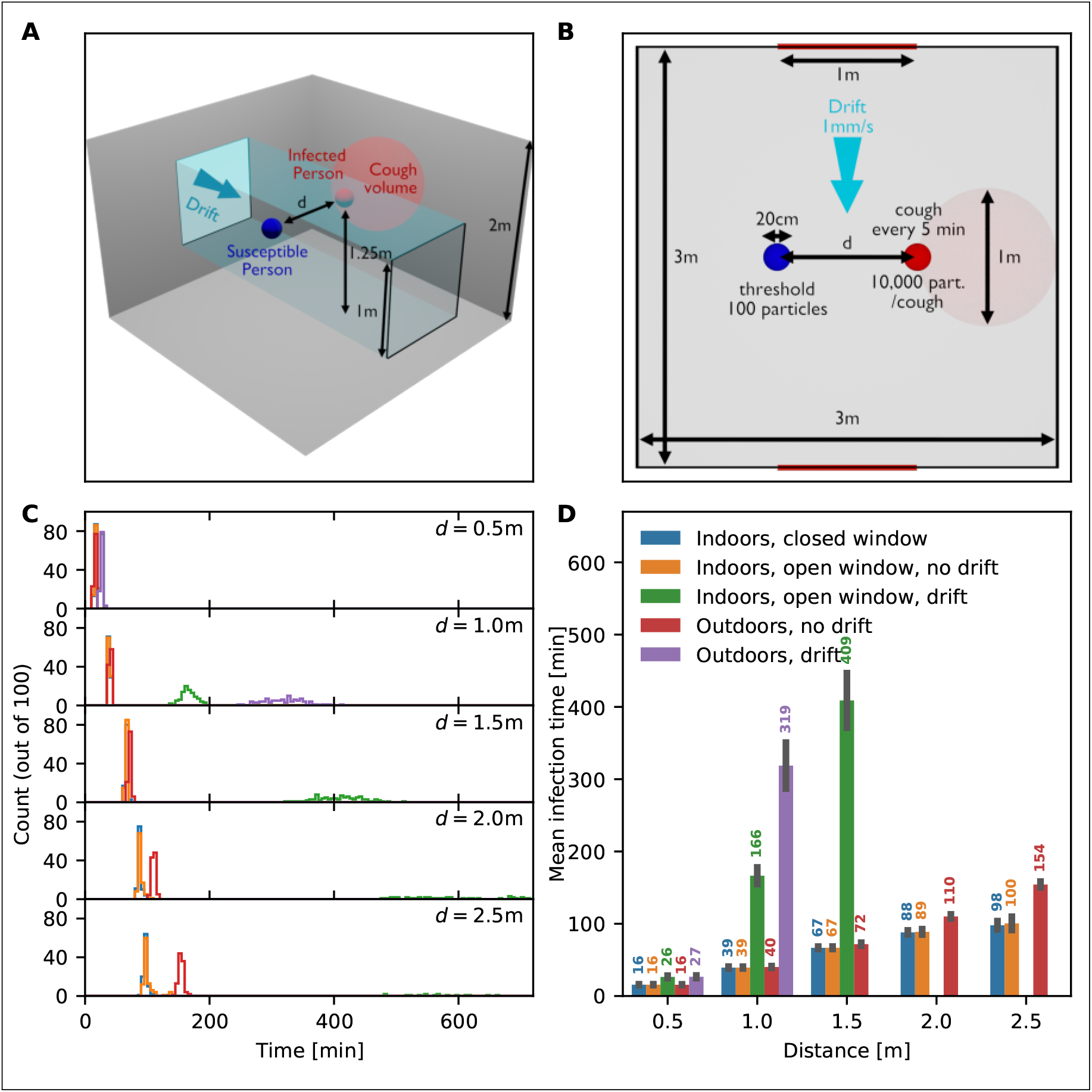
Infection times when an infected person is coughing once every 5 minutes. (A) 3D schematic representation of a closed room containing an infected (red) and a susceptible (blue) person separated by a variable distance *d*. The spherical volume in which 10^4^ coughed particles are dispersed instantly is shaded red. As before, the room contains two windows which can be either open or closed with a possible drift between the windows. (B) Overhead view of (A). Room and window dimensions are displayed in the schematics. (C) Distribution of time of infection for distances *d* = 0.5,1,1.5, 2 and 2.5m and the following scenarios: Indoors with windows closed (blue), indoors with open windows, indoors with open windows and a drift, outdoors without drift (red), and outdoors with drift (purple). (D) Mean infection times for all scenarios. Missing bars indicate scenarios in which at least one realization had an infection time beyond 12 hours.

### 2.4 Effect of wearing a mask

Next, we evaluate the effect of a susceptible person wearing a mask on the time to infection. Surgical masks reduce the viral copy numbers in the fine diffusing fraction by a factor of 2.8 in viral aerosols [10], We run similar simulations as above, where we model a mask by randomly counting only one in 2.8 particles that arrive at the susceptible person on average. When the infected person is breathing only (Fig. 3), the mask increases the time to infection consistently by more than an hour for all distances. At *d* = 0.5*m*, the time to infection is more than doubled, from 95 ± 8 minutes to 200 ± 14 minutes. When the infected person is coughing, the effect of the mask is lessened, but still adds an additional 8 to 28 minutes depending on the distance.

**Figure 3:**
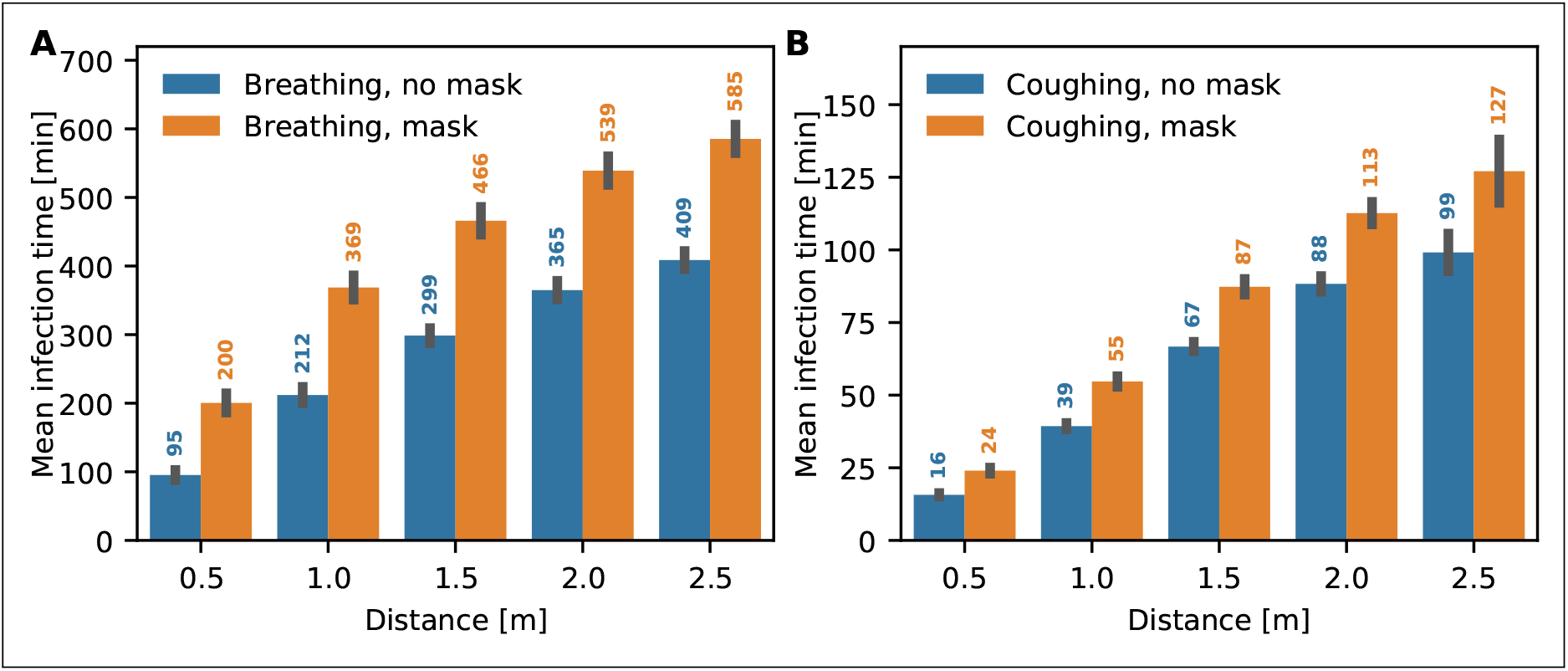
Effect of wearing a mask rejecting on average 2.8th particles for everyone accepted. (A) Mean infection times for various distances *d =* 0.5,1,1.5, 2 and 2.5m (indoors with windows closed), with and without mask when the infected person is breathing only. (B) Mean infection times for the same scenario as in (A), except that the infected person is coughing every 5 minutes.

### 2.5 Parameter sensitivity

Due the large uncertainty in the value of key parameters such as the diffusion coefficient or the threshold of infection, we change their values and explore the consequence on the mean time of infection. Halving the threshold number of particles from 100 to 50 reduces, the time to infection by only between 4 and 13 minutes when the infected person is coughing, depending on the distance (Fig. 4A; see Fig. S3 for the breathing case). Increasing the threshold by an order of magnitude, from 100 to 1000 only roughly doubles the time to infection. Hence, the dependence of time to infection on the threshold is non-linear, as predicted by formula (12). Conversely, the value of diffusion coefficient has a significant effect: an increase from 10^−5^*m*^2^/*s* to 10^−4^*m*^2^/*s* decreases the infection time at *d* = 1*m* from 229 minutes to 39 minutes. Note that due to temperature variation, the change of season from summer to winter leads to a 10% decrease in the diffusion coefficient, which is associated with stronger spread of airborne diseases. Finally, we vary the number of particles emitted during coughing from 5000 to 20000. At a distance of *d* = 1m, this leads to a decrease of infection time from 49 minutes to 32 minutes. Therefore, wide variations in both the threshold number and the number of particles emitted during coughing have a comparatively small effect on the time to infection. In contrast, the diffusion determines the time scale, so its value does directly scale the time to infection. Even though these parameters are not known the trends described above are still valid, if not the exact values of the time to infection.

**Figure 4:**
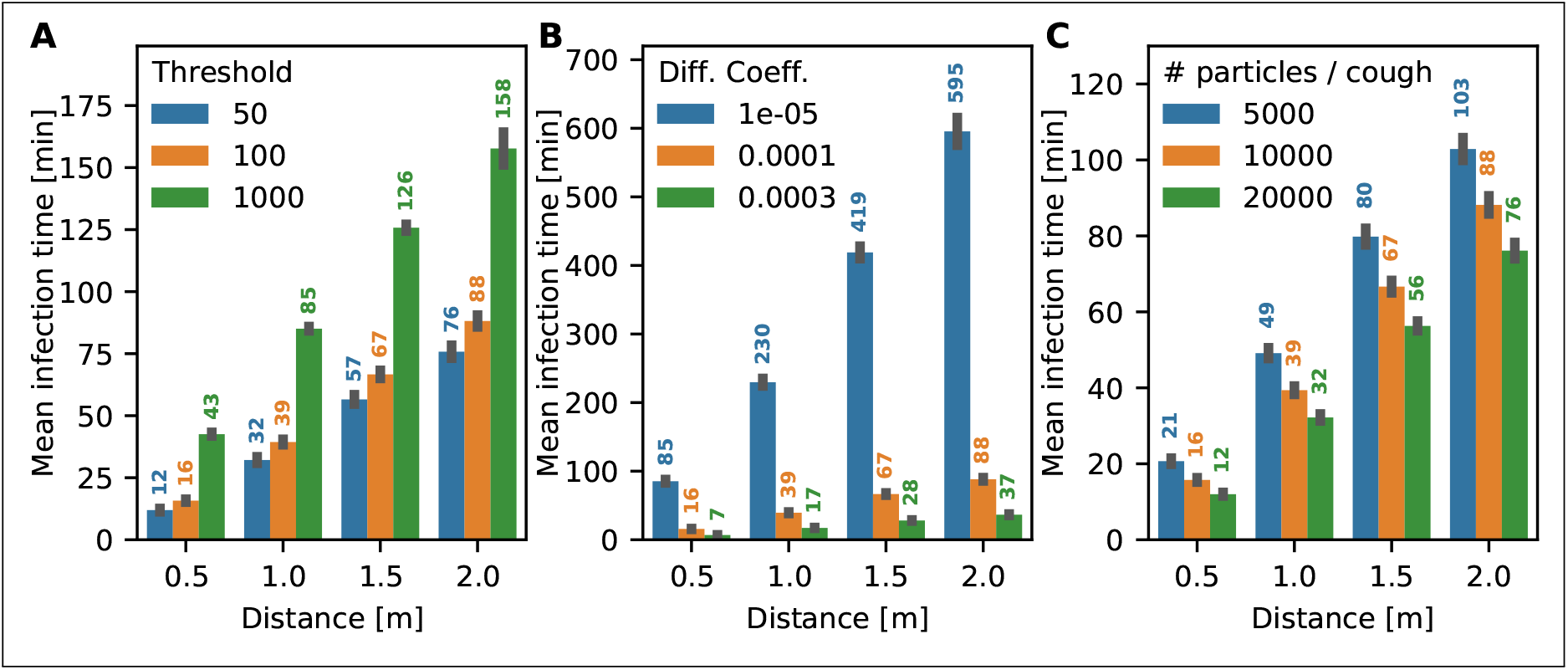
Sensitivity of the mean infection time to the threshold and diffusion coefficient. The mean infection time is estimated for an infected person coughing every 5 minutes in a room with windows closed. (A) Mean infection time for different values of the infection threshold count. (B) Influence of the diffusion coefficient. (C) Mean infection time vs the particle count contained in a cough

## 3 Discussion

Here, we presented a model for airborne viral propagation based on diffusion of aerosols between an infectious and susceptible person [1,2]. We focused on small aerosol particles (< 5*μm*) because they can deeply penetrate into the respiratory tract [4,5], leading to a high probability of infection. A number of parameters used in our simulations, in particular the number of viral particles required for an infection, the number of viral particles emitted during breathing or coughing and the aerosol diffusion coefficient, are not well known and also would be difficult to estimate experimentally. We therefore based their values on other airborne viral illnesses (see sections 4.1 and 4.2). To validate our results, we performed a series of parameter sensitivity studies which show that the trends we observe do not strongly depend on the exact parameter values (Fig. 4).

The most striking result of the present study is the difference in time to infection between the cases when the infected person is breathing normally (e.g. during an asymptomatic infection) to when the infected person is coughing: A susceptible person needs to spend a considerable amount of time in close proximity of a infected person who is not coughing. This picture changes immediately when the infected person is coughing. While we did not look at speaking, we can assume that if the infected person is talking (even if they are not coughing), the picture would be similar to the coughing case.

Coughing three times (once every five minute) lead to infection in 15 minutes. Coughing every 30 seconds roughly halves this time, but does not shorten it by a factor of 10 as could be naively assumed. This is because we assume that the room is completely uncontaminated at the start of the simulation, without any viral particles present, and it takes time for aerosolized viral particles to disperse independently of the coughing frequency. Therefore, this is analogous to a situation where at time zero, the infected person enters the room and starts to cough. While we did not address the reverse situation where the susceptible person enters a room that already contains an infected person (and viral particles have had time to disperse), we can infer that in this case infection will occur much quicker. In a modeling study of influenza transmission, Atkinson and Wein find that a cough leads has a probability of roughly 25% to transmit the disease in a household setting [5,18], which compares well to our findings.

### Distance

Our results suggest that the time to infection increases roughly linearly with distance. Apart from coughing/breathing and ventilation, it is the parameter with the most influence on the infection time.

### Masks

We found that wearing mask can increase the time of infection considerably increases the time it takes to get infected when breathing only, which reinforces that consistent mask wearing reduces asymptomatic transmission. It also increases the time to infection in the coughing case, although the effect is much smaller. Note that we only look at the small aerosol particle fraction for which a mask is less efficient. We assume that the infectious person is coughing in a direction away from the susceptible person which prevents spread via large droplets. Our results here are in line with recent findings that masks can stagger the pandemic over longer times, thereby preventing the saturation of intensive care units in hospitals [19],

### Ventilation

We compared the time to infection indoors when windows are closed to when they are open (Figs. 1 and 2). In the absence of ventilation (i.e. a drift), the role of a window is to prevent the accumulation of viral particle in the room by creating a region that effectively absorbs particles. However, we find this is not enough to substantially affect the time to infection. If, in addition, there is ventilation (even a small drift is sufficient), the time to infection increases markedly. This assumes that this air movement is in any direction but from the infected to the susceptible person. In that case, the time to infection is reduced considerably (Fig. S1). Therefore, we can conclude that ventilation is a good prevention strategy but care must be taken about the direction of air movement.

### Infection threshold number

The thresholds number of particles required for an infection had only little effect on the infection time (Fig. 4). This is due to waves of large numbers of particles being released with each cough event. In the regimes relevant for infection, the time to infection is then determined more by when the next wave of particles reaches the susceptible person rather than by how many are absorbed.

In summary, we believe that the present diffusion-based simulation method yields relevant insights into the relative effects of different scenarios, and also provides a good indication of the magnitude of the time to infection. However, we stress that since the simulation results are dependent on parameter values that are not well known and had to be inferred, the exact time to infection predicted by the simulations should not be taken at face value. Nevertheless, as we showed, the trends we present are valid and are in line with other findings in the literature and public guidance.

## 4 Methods

### 4.1 Model and simulations

The microscopic diffusion coefficient of aerosolized particles is estimated using the Einstein-Kolmogorov formula and is below 10^−10^*m*^2^/*s*. However, the dominant process in this case is not microscopic diffusion, but turbulent diffusion, where the effective diffusivity is increased to *D* = 10^−4^*m*^2^/*s*. To simulate the spread of aerosolized viral particles, we use Brownian motion. The position 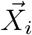 of each particle *i* satisfies the Smoluchowski limit of the Langevin equation [20]

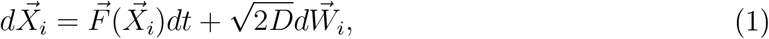

where 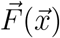 specifies a possible drift due to air movement, *D* is the diffusion coefficient of the aerosolized particle, *W* is a Wiener process. We discretize this equation using the Euler-Maruyama algorithm (forward time stepping).

Interaction with various surfaces is accounted for by various boundary conditions: In the indoors case, particles are reflected by walls, floor and ceiling, placed at the *x,y,z* = 0m and *x, y* = 3m and *z* = 2m positions. Windows are located at the *y* = 0m and *y* = 3m walls, with the extents 0.5*m < z <* 1.5*m* and 1*m* < *x* < 2*m*.

If windows are open, a particle’s trajectory is terminated once it crosses the window boundary (i.e. windows represent absorbing boundaries). In the outdoors case, only the floor at *z* = 0*m* is reflecting and we restrict our computational domain to a spherical domain with a radius of 10*m*. The optional drift is applied with 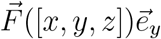 either throughout the domain in the outdoors case, and limited to the region 0.5*m* < *z* < 1.5*m* and 1*m* < *x* < 2*m* in the indoors case (i.e. directly between the windows).

In the case of breathing, the infected person is modelled by a point source that randomly emits particles with an average rate of 5 particles per minute. In the case of coughing, the source of particles is a spherical volume directly in front of the infected person’s head, facing away from the susceptible person and of 1*m* diameter, in which 10^4^ particles are randomly placed every 5 minutes (in the case of coughing).

The susceptible person is modelled as an absorbing spherical surface of 10cm radius, a distance *d* away from the infected person. When a threshold number of particles has been absorbed by the sphere, the time of this event is recorded as the time of infection and the simulation terminated.

### 4.2 Limitations of the method

Our approach, while well-suited to the simulation of the spreading of aerosolized particles, also has limitations when applied to the spread of viral infections:

- **Small vs large droplets:** We only consider the spreading of small particles / droplets. Larger droplets (above *5μm*) are not well described by the diffusion approach. However, these particles will also reasonably quickly drift to the ground due to gravity. Therefore they would modify our results only for small distances between susceptible and infected persons, decreasing the time to infection.
- **Diffusion coefficient:** The diffusion coefficient of the particles is determined by their size. While aerosolized particles have a distribution of sizes, we make the simplification that particles have a fixed size and hence a fixed diffusion coefficient. Since the diffusion coefficient essentially sets the time scale of infection, the value of the diffusion coefficient influences the time to infection. Therefore, the absolute values of the mean time to infection directly depend on the diffusion coefficient, but the qualitative trends (such as the effect of wearing a mask or ventilation) are independent of it.
- **Threshold for infection and coughing:** Some of the parameters are not well-known or need to be inferred from other airborne viral illnesses such as influenza. The threshold number of particles required for an infection is not known for SARS-CoV-2, but medical experts estimate it to be between hundreds and thousands of particles [21,22], We decided to use a threshold number of 100 particles, however the mean time to infection is surprisingly insensitive to the exact threshold (Fig. 4A). The average number of particles emitted during coughing is also not well-known, but similar to the threshold number, the time to infection is not strongly dependent to the exact value (Fig. 4.
- **Walls:** Here walls, the floor and the ceiling are reflective for droplets, but they could also simply stick to surfaces. Removing them would lead to less droplet load, making the mean time to infection longer. However, comparing our indoors and outdoors simulations, this effect is small. We disregard any furniture, or the bodies of the infected and susceptible people, which would likely only have a small effect on the mean time to infection. This is because the threshold is reached by the fastest particle that move directly toward the respiratory tracks.

### 4.3 Deriving the laws of infection based on the threshold hypothesis

Here, we use the theory of diffusion to estimate the flux of droplets to the nose entrance of a susceptible person: after *N*_0_ droplets are instantaneously released following breathing, sneezing or coughing, they diffuse into the surrounding air. We neglect any surrounding surfaces such as floors, walls and, as above, assume that infection occurs when the number of droplets reaches a given threshold *T*. The spatial distribution of droplets satisfies the diffusion equation

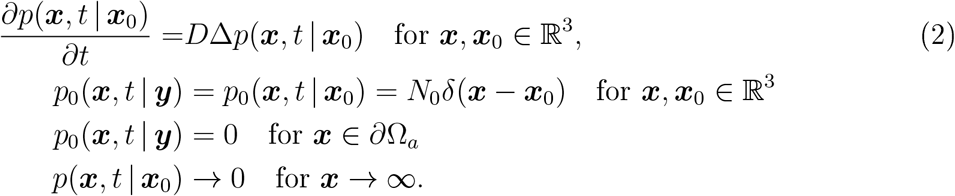

The flux of droplets on an absorbing surface near the face (∂Ω_a_) of a susceptible person is given by

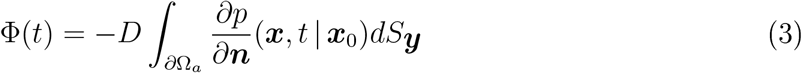

and the cumulative number of particles absorbed is

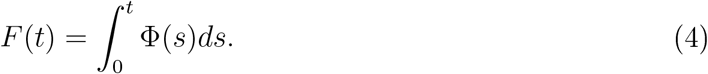

When the source (i.e. the infectious person) is at ***x*** and the target (i.e. the susceptible person) is located at position ***y***, the distribution is given by

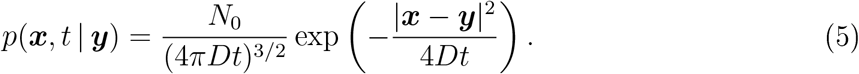

By neglecting the variation of the flux on the surface ∂Ω_a_ when the distance between the two faces is large, δ = |***x*** — ***y***| ≫ 1, the flux can be approximated by

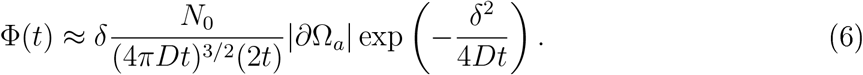

The number of inhaled particles over time is then

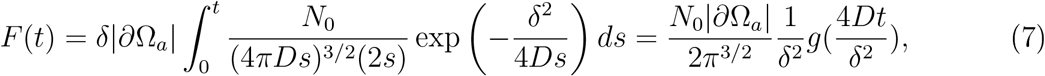

where 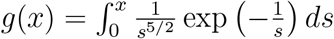. Note that for short times 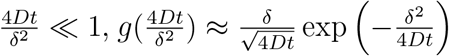 and thus

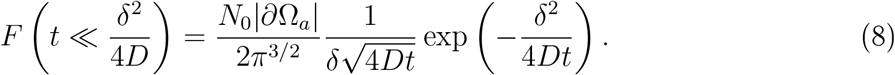

For long times, *g* approaches a constant given by 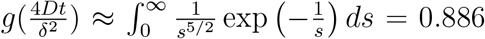 (Fig. SS6). The relevant time scale here is 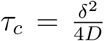, which at δ = 1*m* and with D = 10^−4^m^2^/s, *τ*_*c*_ = 2500*s* ≈ 42*min*. The time of infection *τ*_*inf*_ is achieved when the total number of inhaled particle reaches the threshold *T*_*inf*_ :

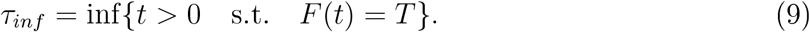

Note that the probability of an infection occurring earlier than *τ*_*inf*_ can be defined by

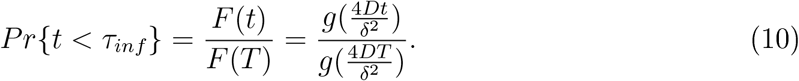

When infection occurs in the time regime before *τ*_*c*_, we can invert relation (8) to get 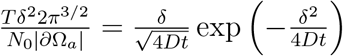 leading to

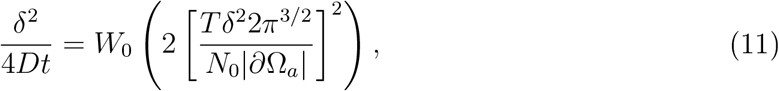

where *W*_0_ is the positive branch of the Lambert function. With W_0_ (*x*) = ln(*x*) – ln(ln *x*) +*o*(1), it can be approximated as

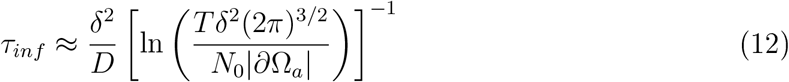

Interestingly, relation (12) shows that the infection time depends weakly on the threshold (as the reciprocal of the logarithm). Expression (12) matches well with data from stochastic simulations for infection times *τ*_*inf*_ < *τ*_*c*_ for a single cough (Fig. S4). Note that we changed the threshold to 2 particles for this, to push the infection time below the *τ*_*c*_ threshold for small distances.

### 4.4 Time distribution of infection for periodic breathing, coughing or sneezing

During repetitive breathing or coughing, we can extend the previous analysis, injecting at the point source a repetitive load of particles. In the periodic case, we can define the elementary injection time Δ*t*_*inj*_ (Δ*t*_*breath*_ or Δ*t*_*Cough*_) leading to condition at the source:

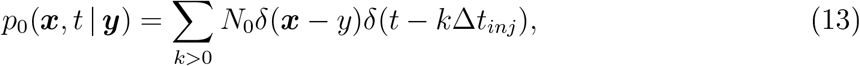

here *N*_0_ is the number of viral particles exhaled by the source person. Using the relations 2 and 7, we obtain the total flux due to breathing only,

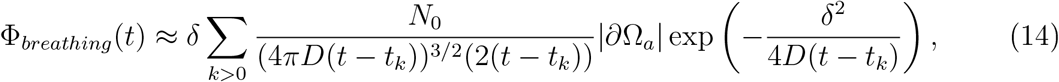

where the sum extend to the duration the susceptible and the infectious persons ar together at distance *δ*. Here *t*_*k*_ = *k*Δ_*breath*_. For coughing, we get the flux

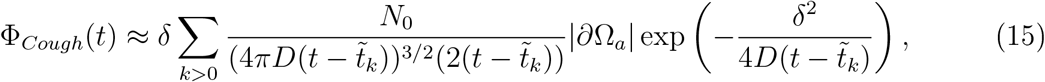

where 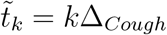. For Breathing and Coughing, we combine the two fluxes:

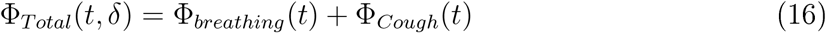

The time of infection is the hist time that the total number of particles reach the threshold *T*_*inf*_

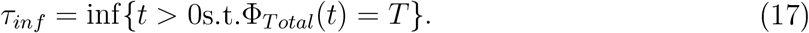

Finally, we can define the probability of infection at time t for distance δ for a viral load threshold T as

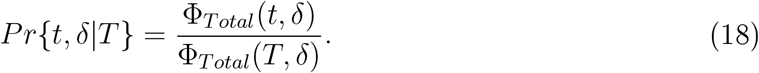

We show that coughing every 30s coughing (500 particles each time) while breathing lead to 4 virus released every 1 min outdoors lead to a rhythmic pattern (Fig. SS5). Note that even after the infectious person leaves the scene, the aerosol concentration decay exponentially.

In the fast load release approximation (in case of very often coughing), by considering that most of viral load is achieved at the maximum of the coughing event, we obtain a linear regime of accumulation:

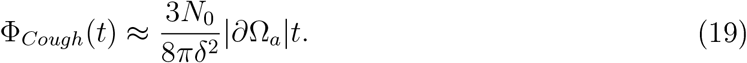

Interestingly, after *Q* viral particles have been released in a ball of radius a and the infectious person has left the room, the law of spreading decays with a rate 1/*t*^3/2^ given by [23] p.257

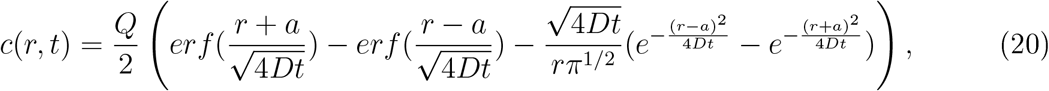

showing a decay in time with

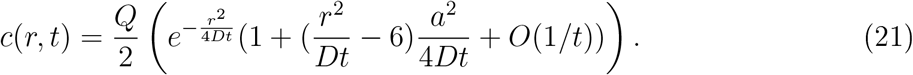

## Data Availability

Manuscript does not contain experimental data.

## 5 Supplementary Figures

**Figure S1:**
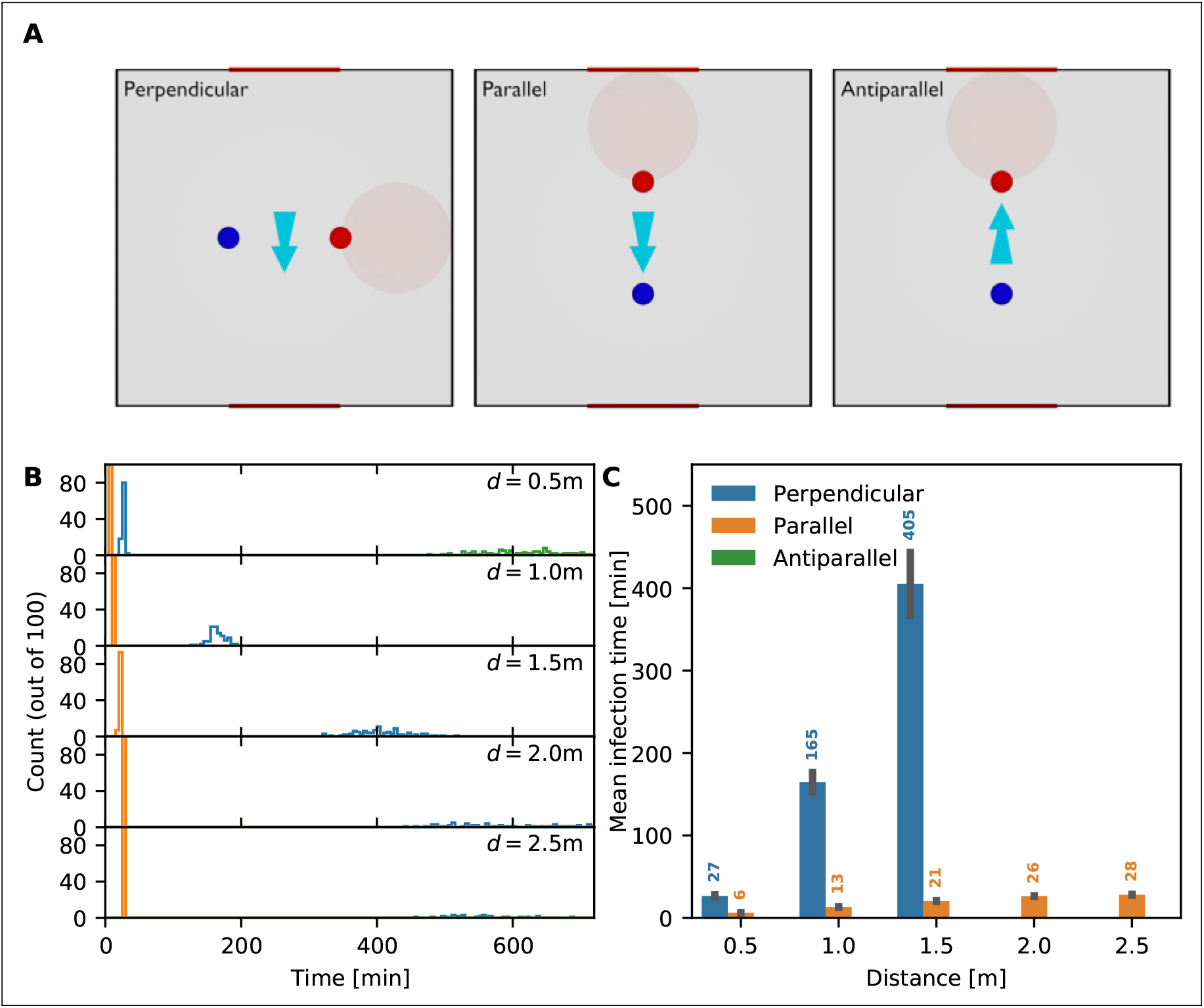
Influence of the direction of air movement. The mean time to infection as a function of distance and air movement direction for an infected person breathing in a room with open windows. (A) The light blue arrow indicates the direction of air movement / ventilation between the two windows. The direction of infection between the infected person (red circle) and the susceptible person (blue circle) can be perpendicular (left panel), parallel (middle panel) and antiparallel (right panel) to the direction of air movement. (B) Distribution of time to infection for distances *d =* 0.5,1,1.5, 2 and 2.5m and the scenarios: perpendicular (blue), parallel (orange) and antiparallel (green). (C) Mean infection time for the different scenarios. Missing bars indicate scenarios in which at least one realization had an infection time beyond 12 hours.

**Figure S2:**
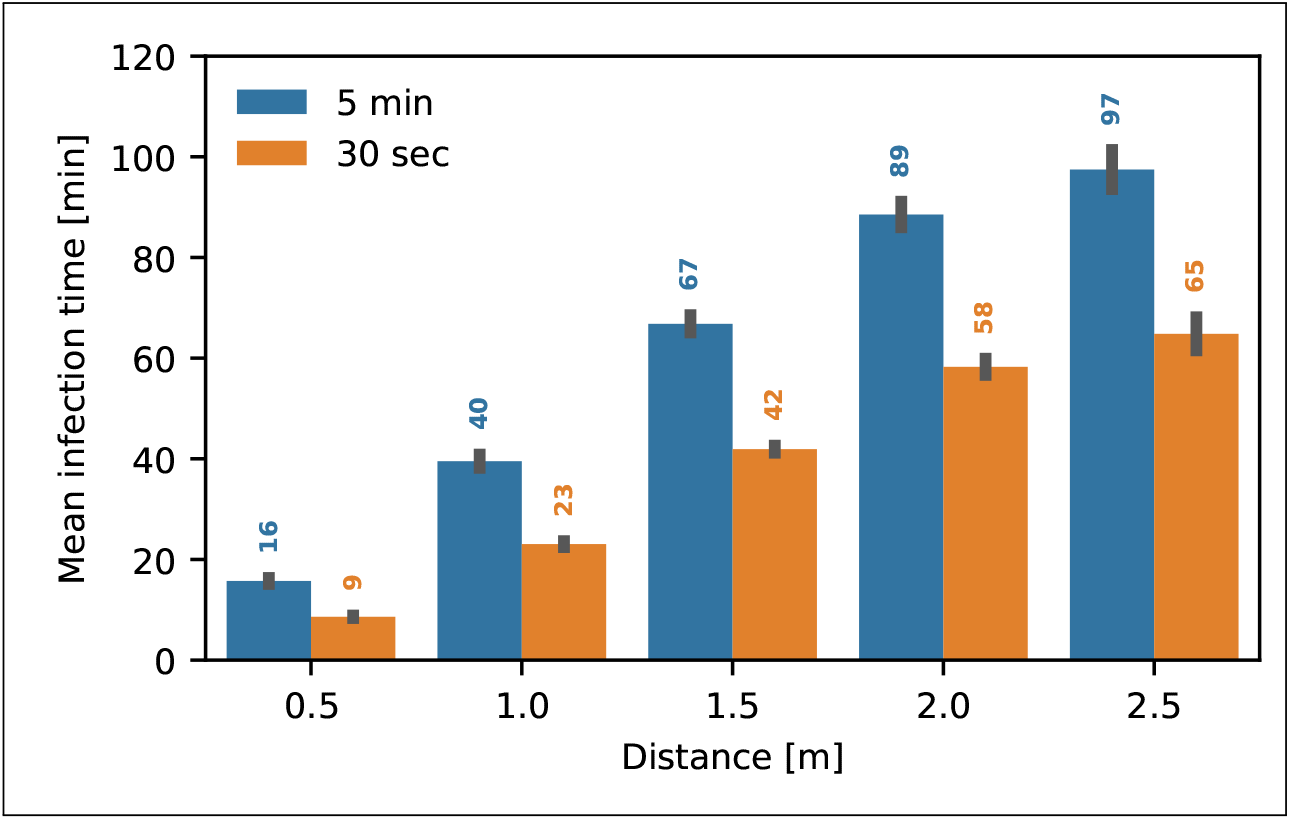
Influence of the cough interval. Mean time to infection for two different cough intervals, 5 minutes (blue) and 30 seconds (orange), and increasing distances *d* between the infected and susceptible persons. Error bars indicate the standard deviation.

**Figure S3:**
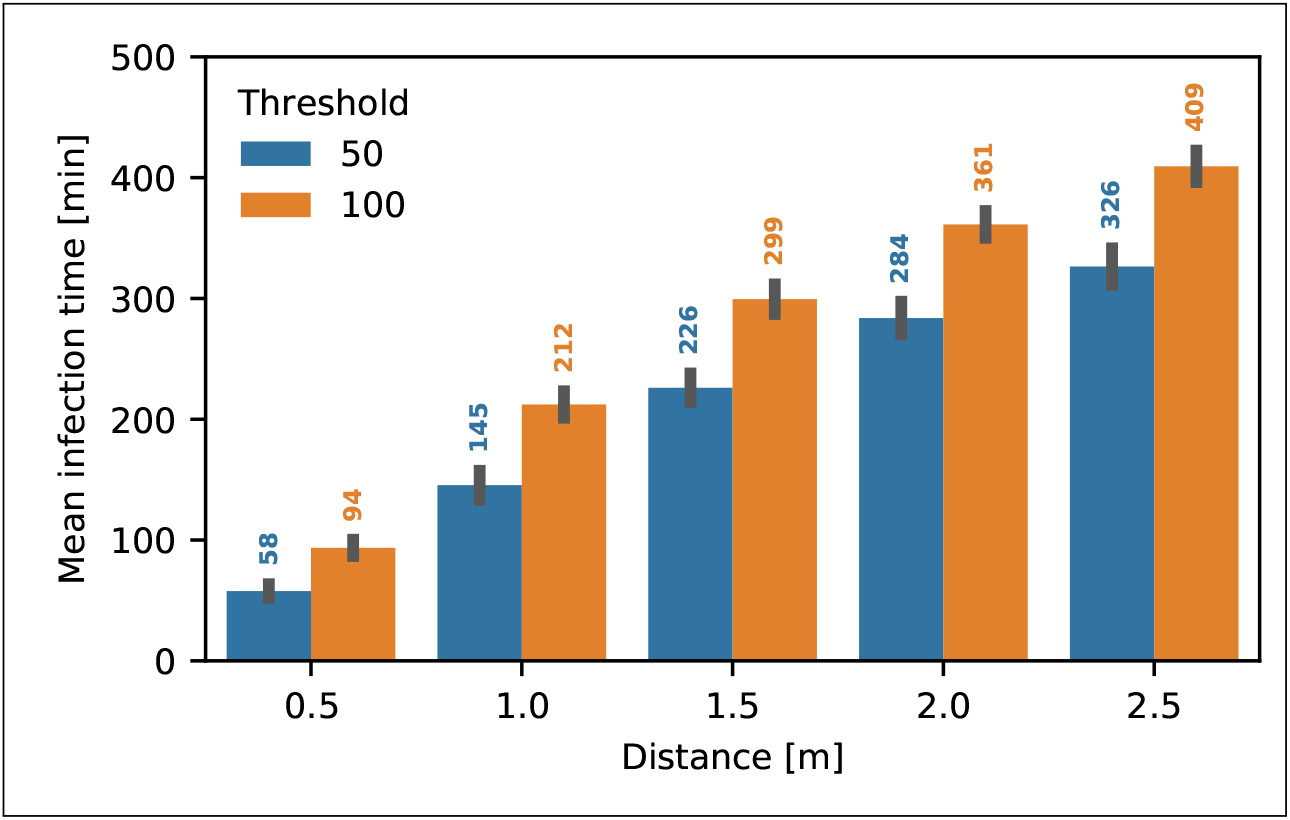
Influence of the threshold number of particles required for infection in the breathing case. Mean time to infection for two different values of the threshold, 50 (blue) and 100 absorbed particles. Error bars indicate the standard deviation.

**Figure S4:**
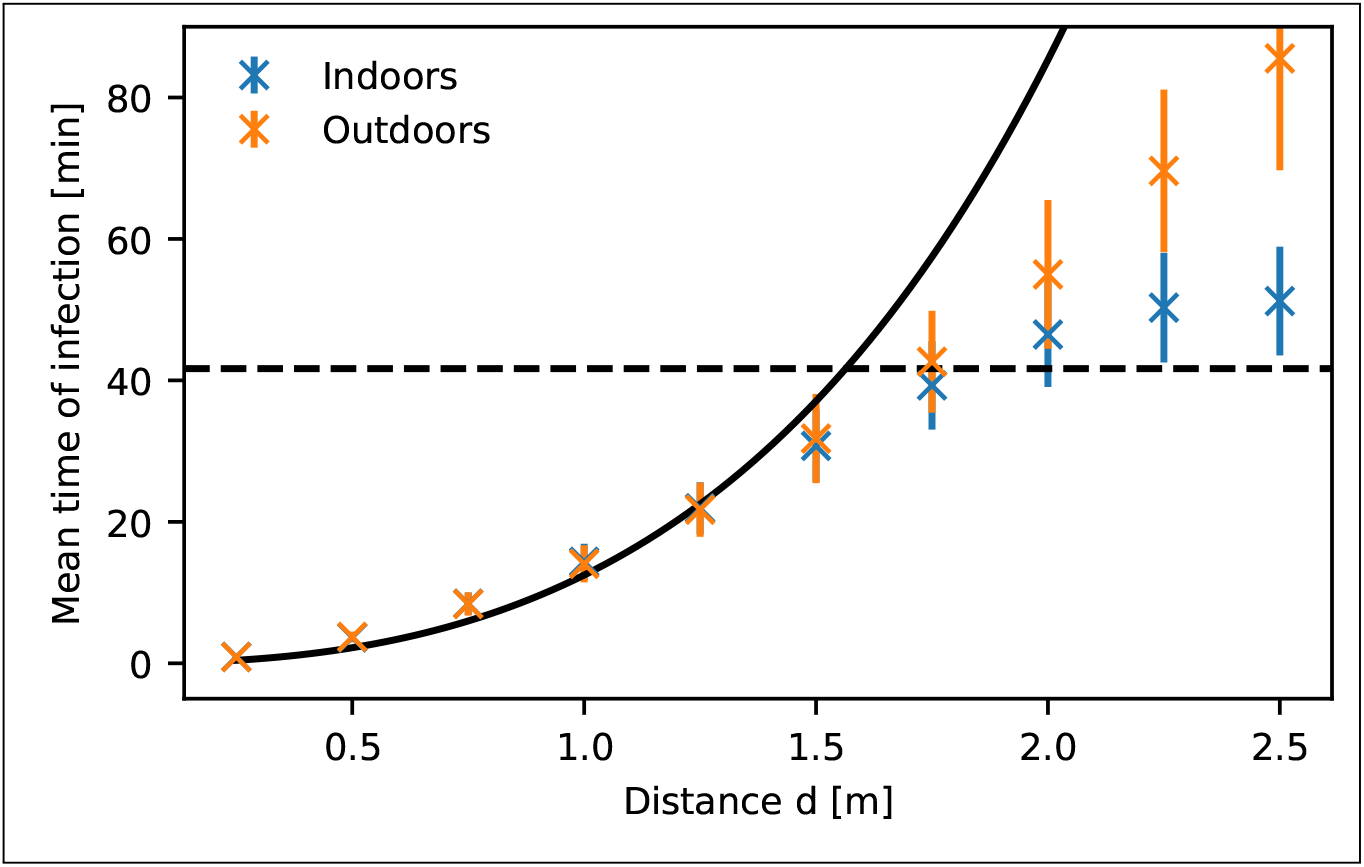
Mean infection time as a function of distance for a single cough. The mean infection time as a function of distance *d* from simulations (crosses) for the indoor case with windows closed (blue) and the outdoor case without any drift (orange) when the infected person is coughing once. Error bars indicate the standard deviation. The solid line shows *τ*_*inf*_ from equation 12, together with the crossover time *τ*_*c*_ (dashed black line). Note that the threshold is set to 2 particles here.

**Figure S5:**
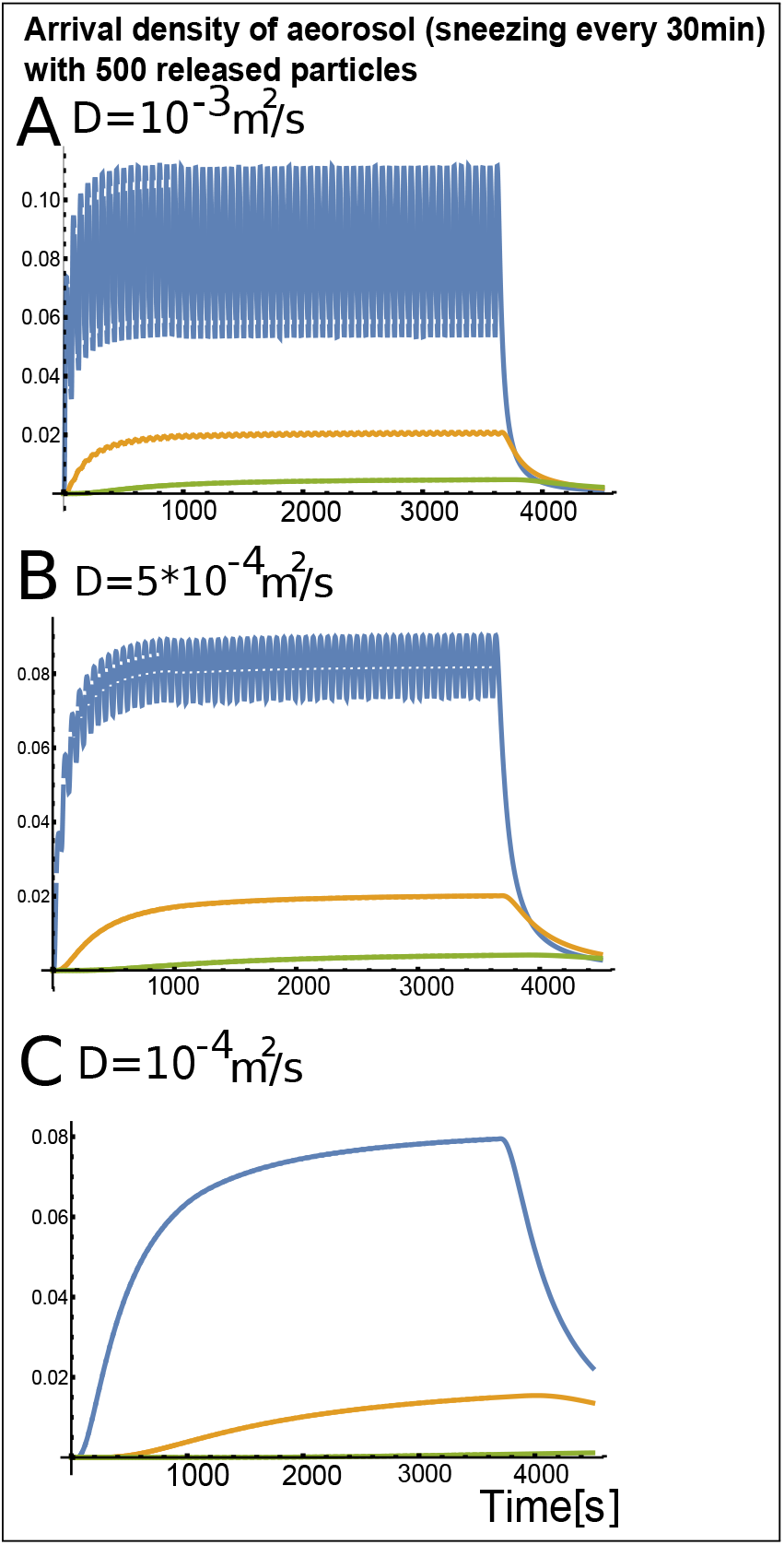
Effect of the diffusion coefficient in a mixed of breathing coughing. Coughing every 20s for 1 hour. After the infectious person leaves the room in three cases and three distances (*δ* = 0.5 (blue), 1 (orange) and 2*m*(green)): (A)*D* = 10^−3^*m*^2^/*s*, (B)*D* = 5.10^−4^*m*^2^/*s* and (C)*D* = 10^−4^*m*^2^*/s*.

**Figure S6:**
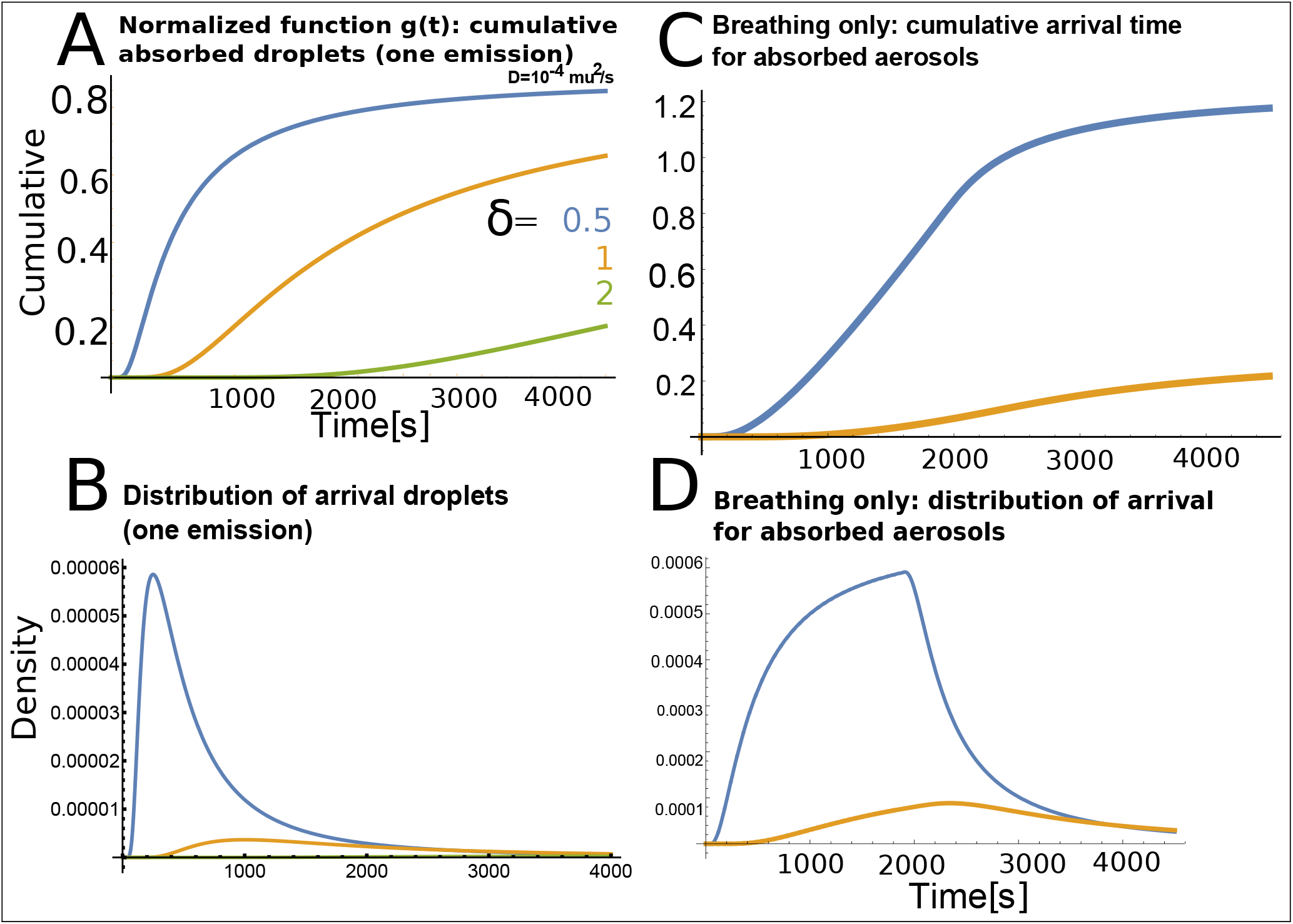
Cumulative and Distribution of viral particles arriving to a target person. Cumulative arrival (A) and distribution (B) of arrival for one event and rhythmic breathing, where 10 viral particles are emitted every minutes during 1800s, after the person leave the room.

## Notes

### Competing Interest Statement

The authors have declared no competing interest.

### Funding Statement

U.D. was supported by a Herchel Smith Postdoctoral Fellowship and acknowledges core funding by
the Wellcome Trust (092096) and CRUK (C6946/A14492). D. H. 's research has received funding from
the European Research Council (ERC) under the European Union's Horizon 2020 research and innovation
programme (grant agreement No 882673), Plan Cancer-INSERM Projet 19CS145-00 and ANR NEUC-0001.

### Author Declarations

nothing to declare

